# Micro-elimination of Hepatitis C virus (HCV) infection in the General Population Cohort in rural Uganda: long-term follow-up to assess feasibility and outcomes of a screening and treatment intervention

**DOI:** 10.1101/2025.10.17.25338164

**Authors:** Joseph Mugisha, Beatrice Kimono, Sheila Lumley, Geraldine O’Hara, Elizabeth Waddilove, Clara Wekesa, George Mgomella, Ronald Makanga, Richard Ndungutse, Moffat Nyirenda, Chris Davis, Emma Thomson, Ponsiano Ocama, Janet Seeley, Philippa C Matthews, Robert Newton

**Affiliations:** Medical Research Council/Uganda Virus Research Institute and London School of Hygiene and Tropical Medicine Uganda Research Unit, Entebbe, Uganda; Peter Medawar Building for Pathogen Research, Nuffield Department of Medicine, University of Oxford, South Parks Road, Oxford OX1 3SY, UK; Department of Infectious Diseases and Microbiology, Oxford University Hospitals NHS Foundation Trust, Oxford, UK; London School of Hygiene and Tropical Medicine, Keppel Street, London WC1E 7HT, UK; The Francis Crick Institute, 1 Midland Road, London NW1 1AT, UK; Makerere University College of Health Sciences in Kampala, Uganda; African Population and Health Research Centre, Kenya; MRC - University of Glasgow Centre for Virus Research, Glasgow, UK; Africa Health Research Institute, KwaZulu Natal, South Africa; Division of Infection and Immunity, University College London, Gower St, London WC1E 6BT, UK; Department of Infection, University College London Hospitals, 235 Euston Rd, London NW1 2BU, UK; Department of Health Sciences, University of York, York, United Kingdom

**Keywords:** HCV, hepatitis, elimination, epidemiology, treatment, DAA, Genotype 4, Africa, Uganda

## Abstract

**Background:** The availability of highly effective curative direct acting antiviral (DAA) therapy for hepatitis C virus (HCV) is a cornerstone of elimination strategies. We report on long-term follow-up as part of a programme that delivered HCV screening and treatment in a population cohort in Uganda.

**Methods:** Screening for HCV, HIV and HBV was offered to >7000 participants in the Kyamulibwa General Population Cohort (GPC) in Kalungu District in rural South-West Uganda in 2011. In 2017, DAA treatment was offered to those individuals who had previously tested HCV RNA positive who could still be traced, with fixed dose combination ledipasvir + sofosbuvir (LED/SOF) for 12 weeks, and post-treatment follow-up at 24 weeks. Clinical review and elastography was repeated in 2023, and verbal autopsy data reviewed.

**Results:** 13 individuals tested HCV RNA positive, of whom five had been born in Uganda and eight originated from Rwanda. The median age at HCV diagnosis was 61 (range 48-90) and 10/13 (77%) were male. Six years later, five had died, one had left the area, and seven individuals were traced, all of whom accepted treatment, with confirmed cure (sustained virologic suppression (SVR)). After a further six year interval, four of those treated were followed up. Among those who had died, a high prevalence of liver disease was suggested by verbal autopsies.

**Conclusion:** Among individuals offered DAA treatment, acceptance and cure rate were high. In this setting, HCV infection likely contributed to mortality, and affected older adults and migrants, suggesting these groups might be priorities for future micro-elimination programmes.

## INTRODUCTION

Global goals have been set for the elimination of Hepatitis C virus (HCV) as a public health threat by 2030 [1]. Oral direct acting antiviral (DAA) treatment for HCV infection is safe, well-tolerated and has high cure rates with sustained virological response (SVR) of >98%, but a 2024 report published by the World Health Organization (WHO) highlights that we are not on track to meet elimination targets in many populations [2], reflecting challenges particularly for resource-limited settings [3]. For many people living in Africa, there is inadequate access to robust HCV diagnostic testing [4], while for those diagnosed, there may be no pathway to affordable treatment [2]. HCV genotypes (Gt) prevalent in some African populations responded less well to original first line DAA combinations (e.g. Gt-4r [5]), due to the higher prevalence of viral polymorphisms associated with resistance. Newer pangenotypic regimens have increased cure rates, although some viral variants have been associated with lower treatment success [6].

Despite the challenges, there are now examples of highly successful African HCV programmes (e.g. in Rwanda [7], Egypt [8] and Nigeria [9]), and ‘micro-elimination’ projects have shown success in targeting defined groups [2]. Initiatives have been resourced by the Global Fund, and supported by the Clinton Health Access Initiative (CHAI) to negotiate quality-assured and affordable generic medications. Uganda is one of 38 ‘focus countries’ listed in a World Health Organization (WHO) report, which together account for ∽80% of viral hepatitis infections and deaths [2]. We here report 12-year follow-up of adults diagnosed with HCV infection in rural Uganda, to explore challenges, feasibility, and long-term outcomes.

## METHODS

### Population setting

This project was based in the Kyamulibwa General Population Cohort (GPC) in rural South-West Uganda, with recruitment and laboratory tests conducted as previously described [10,11]. In 2011, screening for HCV, HIV and HBV was offered to participants as a component of a routine clinical survey [12], and HCV sequencing was undertaken in those with detectable viraemia [11]. The study timeline is presented in **Figure 1**.

**Figure 1.**
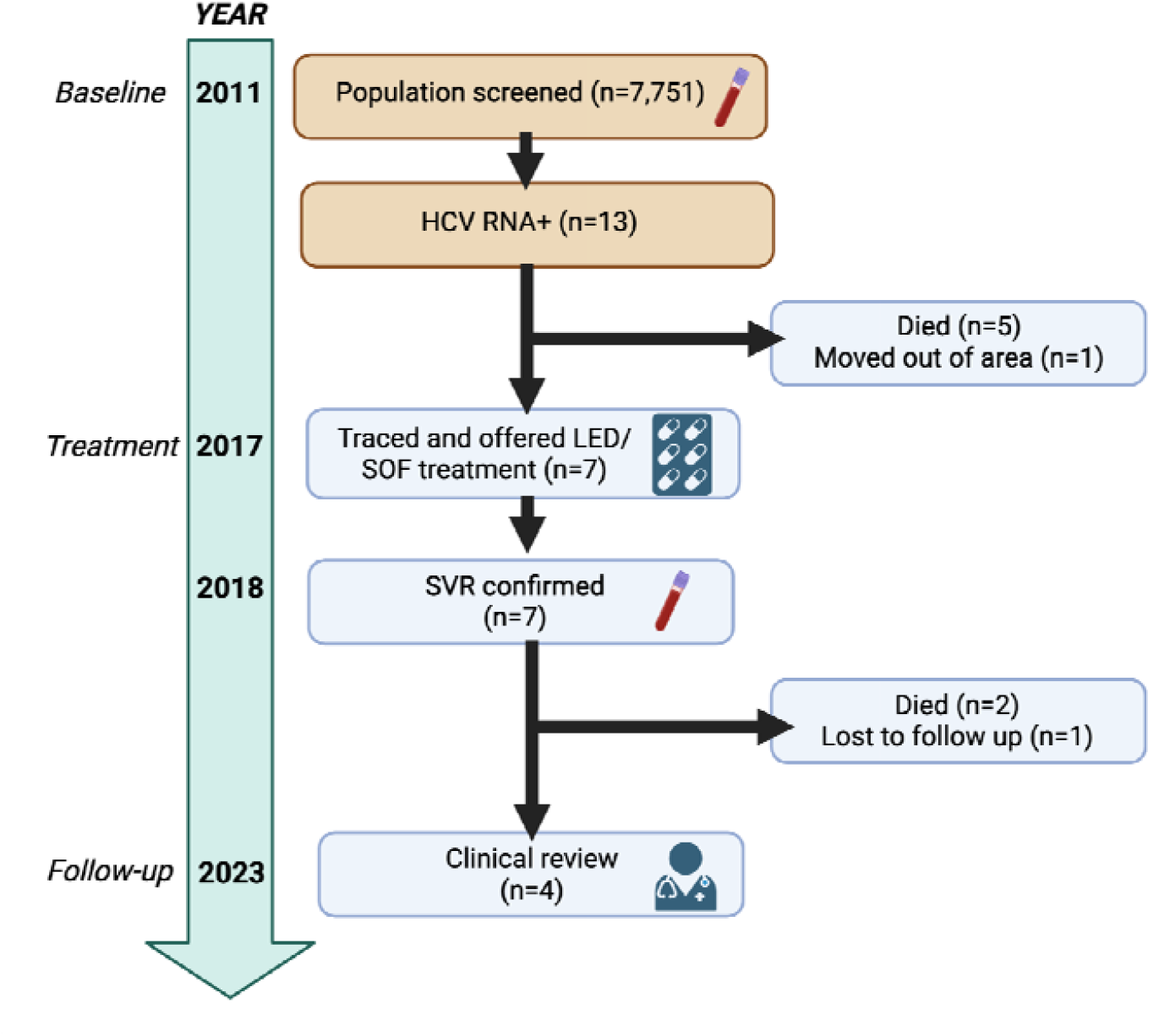
Timeline showing diagnosis, treatment and follow-up of adults with HCV infection in the Kyamulibwa General Population cohort (GPC) in Kalungu District in Southern Uganda. Orange boxes highlight parts of the study previously reported [11].

### HCV treatment protocol

In 2017, when DAA treatment could be accessed, individuals who had tested HCV RNA positive and could be traced were offered treatment with a daily fixed-dose combination of 90mg ledipasvir (LED) and 400mg sofosbuvir (SOF) for 12 weeks, based on contemporaneous guidelines [13–15] (**S1**). SVR was defined as undetectable HCV RNA at 12-24 weeks after the end of treatment. Liver stiffness was measured by transient elastography (TE) using Fibroscan™ (Echosens, Paris, France) at diagnosis, at treatment initiation, and at follow-up in 2023. Details of our training and feedback are reported [16]. For those who died, verbal autopsies were reviewed when available, based on previously reported methods [17].

### Ethics

Ethical approval for the GPC was granted by the UVRI Research and Ethics Committee (GC/127/710) and the Uganda National Council for Science and Technology (SS 4981), and participants provided written informed consent. HCV treatment was prescribed and managed on clinical grounds. Follow up in 2023 was undertaken by the Uganda Liver Disease Study (‘ULiDS’) with ethics approval from the University of Oxford Research Ethics Committee (OxTREC), ref 50-18.

## RESULTS

### Diagnosis and treatment of individuals living with HCV infection

Active HCV infection was identified in 13 individuals as previously reported [11] (**Table 1** and **S2**). At diagnosis, none had cirrhosis or decompensated liver disease. Between 2011 and 2017, five individuals died and one left the area. Seven remaining individuals were contacted and offered treatment, all of whom accepted 12 weeks of DAA therapy. At treatment initiation, liver enzymes were within the normal range (median ALT 26 U/L, **Table 1**) and median TE measurement was 6.3kPa, although two individuals had progressed to meet elastography criteria for cirrhosis (TE 13.3kPa and 14.1kPa), one of whom was also living with HIV. No side-effects were reported during weekly follow-up on treatment.

**Table 1:**
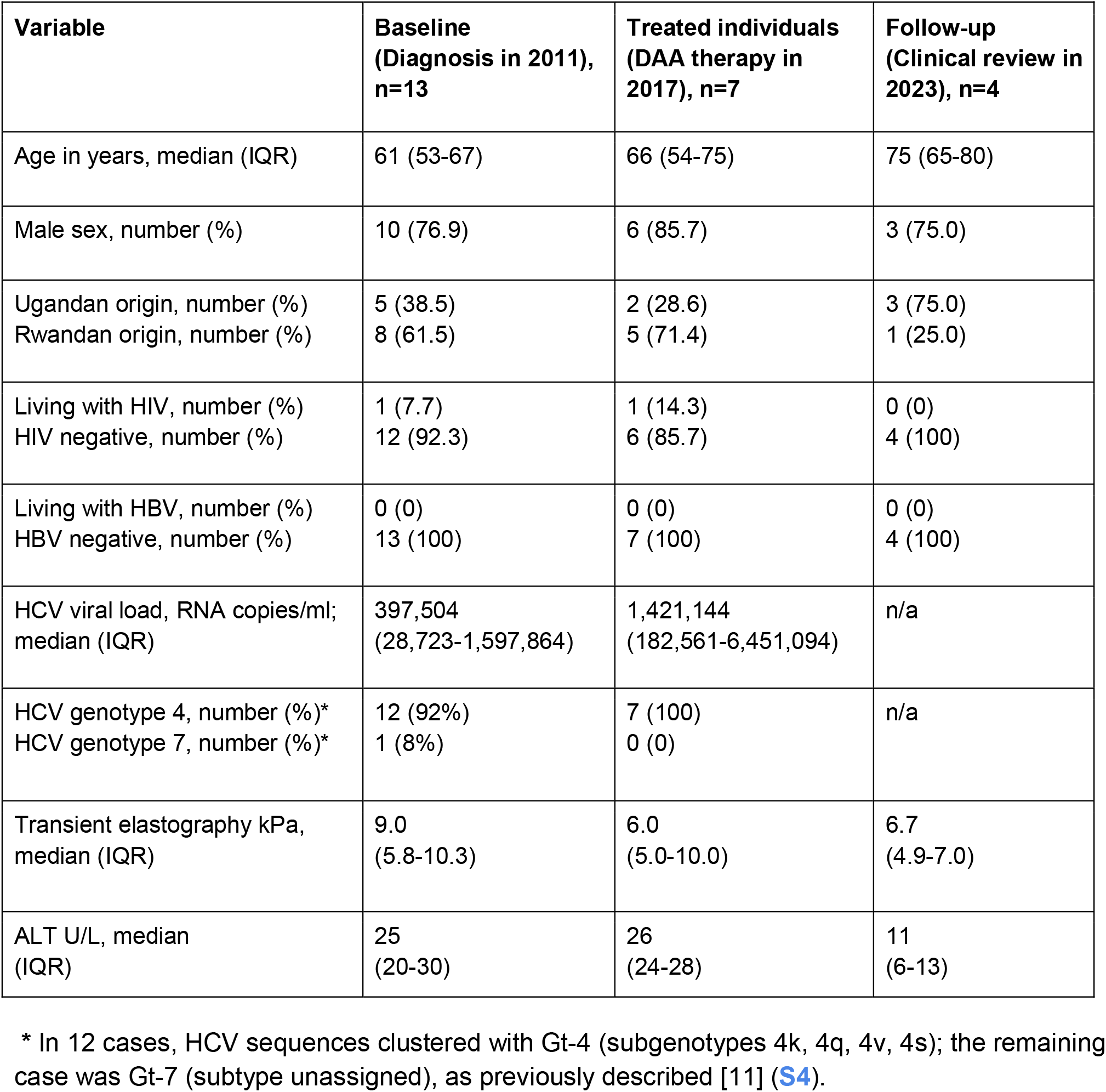
Characteristics of individuals living with HCV infection in the Ugandan General Population Cohort (GPC), at baseline, treatment and follow-up timepoints.

| **Variable** | **Baseline**<br>**(Diagnosis in 2011),**<br>**n=13** | **Treated individuals**<br>**(DAA therapy in**<br>**2017), n=7** | **Follow-up**<br>**(Clinical review in**<br>**2023), n=4** |
| --- | --- | --- | --- |
| Age in years, median (IQR) | 61 (53-67) | 66 (54-75) | 75 (65-80) |
| Male sex, number (%) | 10 (76.9) | 6 (85.7) | 3 (75.0) |
| Ugandan origin, number (%)<br>Rwandan origin, number (%) | 5 (38.5)<br>8 (61.5) | 2 (28.6)<br>5 (71.4) | 3 (75.0)<br>1 (25.0) |
| Living with HIV, number (%)<br>HIV negative, number (%) | 1 (7.7)<br>12 (92.3) | 1 (14.3)<br>6 (85.7) | 0 (0)<br>4 (100) |
| Living with HBV, number (%)<br>HBV negative, number (%) | 0 (0)<br>13 (100) | 0 (0)<br>7 (100) | 0 (0)<br>4 (100) |
| HCV viral load, RNA copies/ml;<br>median (IQR) | 397,504<br>(28,723-1,597,864) | 1,421,144<br>(182,561-6,451,094) | n/a |
| HCV genotype 4, number (%)*<br>HCV genotype 7, number (%)* | 12 (92%)<br>1 (8%) | 7 (100)<br>0 (0) | n/a |
| Transient elastography kPa,<br>median (IQR) | 9.0<br>(5.8-10.3) | 6.0<br>(5.0-10.0) | 6.7<br>(4.9-7.0) |
| ALT U/L, median<br>(IQR) | 25<br>(20-30) | 26<br>(24-28) | 11<br>(6-13) |

\* In 12 cases, HCV sequences clustered with Gt-4 (subgenotypes 4k, 4q, 4v, 4s); the remaining case was Gt-7 (subtype unassigned), as previously described [11] (S4).

### Follow-up

SVR was confirmed at 12-24 weeks post-treatment in all treated individuals. In addition to five deaths preceding HCV treatment, a further two individuals died after receiving HCV treatment. Verbal autopsy data were available in six cases (**S3**), of whom four had a reported history compatible with liver disease (abdominal pain/swelling, bleeding, jaundice, encephalopathy,peripheral oedema), and a further individual met TE criteria for cirrhosis. Four individuals attended review in 2023, with normal TE scores and ALT.

## DISCUSSION

In this rural Ugandan setting, treatment was initially delayed by lack of access to DAA medication, and only 7/13 individuals could be offered treatment. However, all of these accepted treatment and achieved SVR, highlighting the feasibility of HCV micro-elimination based on community-based testing and treatment (**Table 2**). Clinical records and verbal autopsies suggest liver disease as a contributor to the majority of deaths; this is likely related to HCV infection, but other causes of liver disease including alcohol excess are also known to be common on this population[12,18], and this is a vulnerable group of older adults who may have been living with other comorbidities. The drop in median fibrosis score between initial diagnosis and starting treatment highlights that those with higher fibrosis scores died before treatment became available.

**Table 2:**
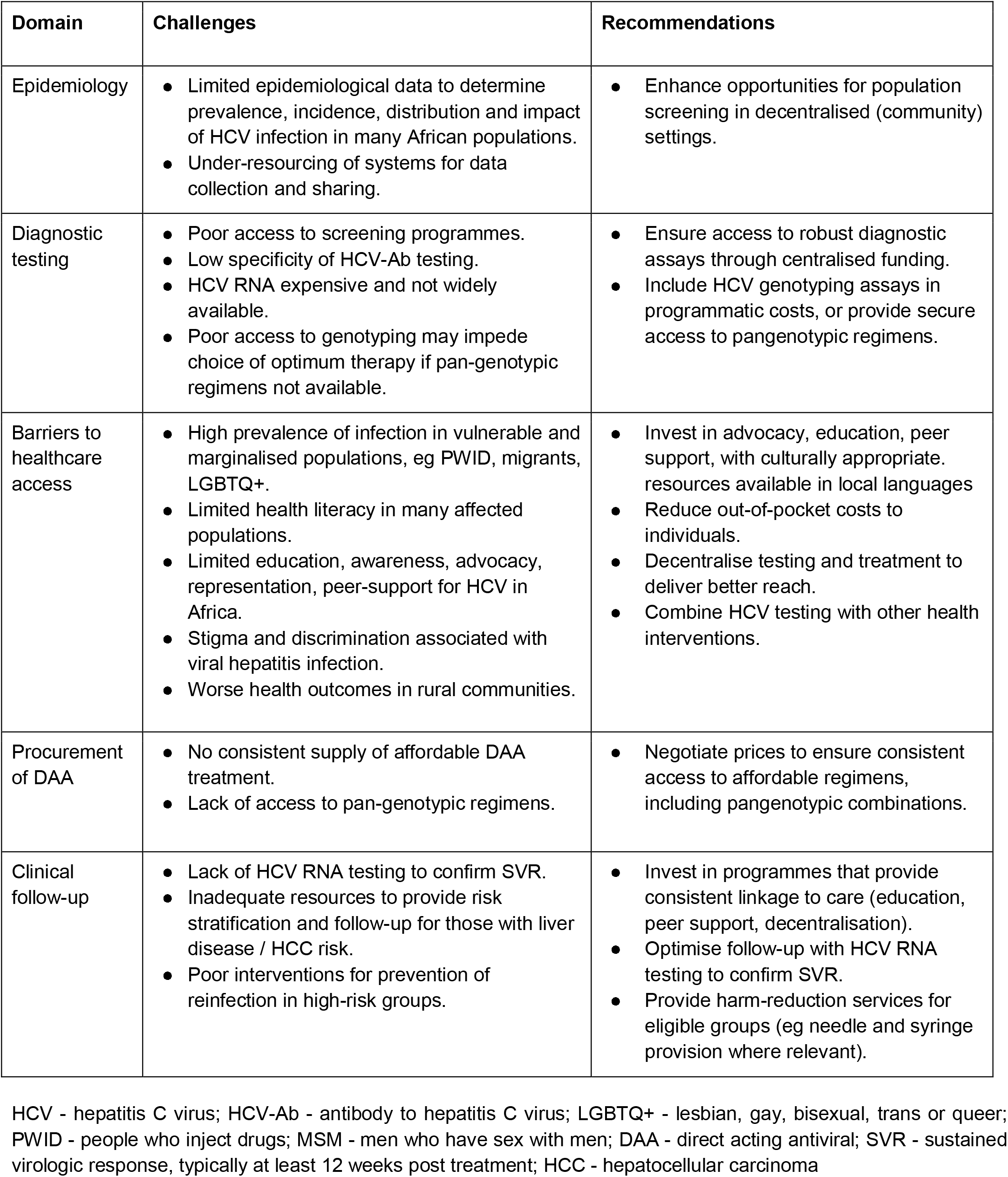
Addressing challenges for HCV micro-elimination in populations in the WHO African region.

In the global journey towards HCV elimination, some individuals and communities have been labelled as ‘difficult to reach’ and genotypes as ‘difficult to treat’ [5]. Overturning these paradigms and delivering equitable interventions demands decentralisation of health services to reach diverse and marginalised communities, and provision of accessible and affordable pangenotypic therapy (e.g. SOF + daclatasvir (DAC)) [19,20]. Micro-elimination is feasible [21], and SOF-based treatment is acceptable, safe and effective [22]. Encouragingly, a study evaluating a minimal monitoring approach with 12 weeks of SOF + velpatasvir (VEL) in the absence of pre-treatment genotyping was safe and achieved SVR rates comparable to settings implementing standard monitoring [23]. Uganda has now committed domestic resources to the viral hepatitis response, and has registered the SOF/DAC fixed dose combination[2]. Cost analyses (including data from Uganda) predict that for every US $1 invested, return on investment will be US $1.7-3.3 based on direct costs averted for the healthcare system [24]. However, many countries are still paying above the benchmark cost for DAA therapy [2].

Older adults in African communities may be more at risk of HCV infection [25], for example as a result of nosocomial transmission (particularly contaminated needles during past vaccination campaigns) or scarification practices [26,27]. Risks have reduced over time due to improved protocols in healthcare protocols for prevention of infection transmission, and shifts in cultural customs. Individuals of Rwandan origin may have moved to Uganda many years before, possibly at times of food shortage, and HCV infection may have occurred at some point in their childhood in Rwanda [28]. Close partnerships with communities are needed in order to provide information, promote testing and support access to harm reduction services where relevant,while respecting cultural and religious beliefs, and recognising the potential impact of stigma and discrimination.

There are significant regional variations in HCV prevalence in African populations [29]. We only identified a small number of individuals living with HCV, and further data are needed to represent different sub-populations including urban dwellers, and regions with higher rates of migrancy (particularly adjoining Uganda’s borders with Rwanda and South Sudan). Individuals who are most at risk of HCV infection may experience barriers to testing and treatment, and we are unable to exclude the possibility that some local cases of infection were overlooked.

Progress towards HCV elimination in Uganda is feasible, using pangenotypic DAA therapy delivered through decentralised programmes with minimal monitoring. However, enhanced data collection, ongoing commitment to resource allocation, and consistent intervention with affordable and accessible testing, treatment and follow-up is needed, especially to reduce inequities in vulnerable and marginalised groups.

## Data Availability

All data reported are included in the supplementary file.

## ACKNOWLEDGEMENTS

We are grateful to other members of the UVRI team and collaborators who were involved in the original HCV data collection and analysis, including Clara Wekesa, Gershim Asiki, Anatoli Kamali, Brian Ssengendo, and Moses Kwizera Mbonye.

## REFERENCES

[1] Hepatitis, (n.d.). https://www.who.int/health-topics/hepatitis/elimination-of-hepatitis-by-2030 (accessed July 6, 2024).

[2] World Health Organization Global Hepatitis Report 2024: Action for Access in Low- and Middle-Income Countries. https://www.who.int/publications/i/item/9789240091672. ISBN: 978-92-4-009167-2, (n.d.).

[3] S. Lanini, P.J. Easterbrook, A. Zumla, G. Ippolito, Hepatitis C: global epidemiology and strategies for control, Clin. Microbiol. Infect. 22 (2016) 833–838.

[4] G.J. Dore, J.J. Feld, Hepatitis C virus therapeutic development: in pursuit of “perfectovir,” Clin. Infect. Dis. 60 (2015) 1829–1836.

[5] F. Shumbusho, A.F. Liu, F. Kateera, J. Kabahizi, S. Nsanzaimana, J. Serumondo, J. Damascene Makuza, P.M. Grant, E. Musabeyezu, C. Muvunyi, N. Gupta, Risk factors for difficult-to-treat hepatitis C virus genotype 4r in Rwanda and implications for elimination in sub-Saharan Africa, J. Viral Hepat. 28 (2021) 682–686.

[6] J. Dietz, C. Graf, C.P. Berg, K. Port, K. Deterding, P. Buggisch, K.-H. Peiffer, J. Vermehren, G. Dultz, A. Geier, F.P. Reiter, T. Bruns, J.M. Schattenberg, E. Durmashkina, T. Gustot, C. Moreno, J. Trauth, T. Discher, J. Fischer, T. Berg, A.E. Kremer, B. Müllhaupt, S. Zeuzem, C. Sarrazin, European HCV Resistance Study Group, Rare HCV subtypes and retreatment outcomes in a cohort of European DAA-experienced patients, JHEP Rep. 6 (2024) 101072.

[7] M.P. Nisingizwe, J.D. Makuza, N.Z. Janjua, N. Bansback, B. Hedt-Gauthier, J. Serumondo, E. Remera, M.R. Law, The Cascade of Care for Hepatitis C Treatment in Rwanda: A Retrospective Cohort Study of the 2017-2019 Mass Screening and Treatment Campaign, Viruses 15 (2023). 10.3390/v15030661.

[8] I. Waked, G. Esmat, A. Elsharkawy, M. El-Serafy, W. Abdel-Razek, R. Ghalab, G. Elshishiney, A. Salah, S. Abdel Megid, K. Kabil, M.H. El-Sayed, H. Dabbous, Y. El Shazly, M. Abo Sliman, K. Abou Hashem, S. Abdel Gawad, N. El Nahas, A. El Sobky, S. El Sonbaty, H. El Tabakh, E. Emad, H. Gemeah, A. Hashem, M. Hassany, N. Hefnawy, A.N. Hemida, A. Khadary, K. Labib, F. Mahmoud, S. Mamoun, T. Marei, S. Mekky, A. Meshref, A. Othman, O. Ragab, E. Ramadan, A. Rehan, T. Saad, R. Saeed, M. Sharshar, H. Shawky, M. Shawky, W. Shehata, H. Soror, M. Taha, M. Talha, A. Tealaab, M. Zein, A. Hashish, A. Cordie, Y. Omar, E. Kamal, I. Ammar, M. AbdAlla, W. El Akel, W. Doss, H. Zaid, Screening and Treatment Program to Eliminate Hepatitis C in Egypt, N. Engl. J. Med. 382 (2020) 1166–1174.

[9] C.E. Boeke, C. Adesigbin, C. Agwuocha, A. Anartati, H.T. Aung, K.S. Aung, G.S. Grover, D. Ngo, E. Okamoto, A. Ngwije, S. Nsanzimana, S. Sindhwani, G. Singh, L.P. Sun, N. Van Kinh, W. Waworuntu, C. McClure, Initial success from a public health approach to hepatitis C testing, treatment and cure in seven countries: the road to elimination, BMJ Glob Health 5 (2020). 10.1136/bmjgh-2020-003767.

[10] G. Asiki, G. Murphy, J. Nakiyingi-Miiro, J. Seeley, R.N. Nsubuga, A. Karabarinde, L. Waswa, S. Biraro, I. Kasamba, C. Pomilla, D. Maher, E.H. Young, A. Kamali, M.S. Sandhu, G. P. C. team, The general population cohort in rural south-western Uganda: a platform for communicable and non-communicable disease studies, Int. J. Epidemiol. 42 (2013) 129– 141.

[11] C. Davis, G.S. Mgomella, A. da Silva Filipe, E.H. Frost, G. Giroux, J. Hughes, C. Hogan, P. Kaleebu, G. Asiki, J. McLauchlan, M. Niebel, P. Ocama, C. Pomila, O.G. Pybus, J. Pépin, P. Simmonds, J.B. Singer, V.B. Sreenu, C. Wekesa, E.H. Young, D.G. Murphy, M. Sandhu, E.C. Thomson, Highly diverse hepatitis C strains detected in sub-Saharan Africa have unknown susceptibility to direct-acting antiviral treatments, Hepatology 69 (2019) 1426– 1441.

[12] C. Campbell, J. Mugisha, B. Kimono, E. Waddilove, R. Makanga, T. Wang, F.N. Muzaale, P.C. Matthews, R. Newton, Liver, cardiovascular and metabolic factors as predictors of allcause mortality in a rural Ugandan Cohort, medRxiv (2025). 10.1101/2025.04.03.25325170.

[13] AASLD/IDSA HCV Guidance Panel, Hepatitis C guidance: AASLD-IDSA recommendations for testing, managing, and treating adults infected with hepatitis C virus, Hepatology 62 (2015) 932–954.

[14] I.M. Jacobson, The HCV Treatment Revolution Continues: Resistance Considerations, Pangenotypic Efficacy, and Advances in Challenging Populations, Gastroenterol. Hepatol. 12 (2016) 1–11.

[15] A. Kohli, R. Kapoor, Z. Sims, A. Nelson, S. Sidharthan, B. Lam, R. Silk, C. Kotb, C. Gross, G. Teferi, K. Sugarman, P.S. Pang, A. Osinusi, M.A. Polis, V. Rustgi, H. Masur, S. Kottilil, Ledipasvir and sofosbuvir for hepatitis C genotype 4: a proof-of-concept, single-centre, open-label phase 2a cohort study, Lancet Infect. Dis. 15 (2015) 1049–1054.

[16] M. Anderson, G. Sukali, J.-L. Upton, E. Waddilove, L. Downs, S. Lumley, M. Delphin, B. Kimono, J. Mugisha, J. Seeley, T. Khoza, S. Temmers, R. Newton, N. Mhlongo, V. Naidoo, P. Matthews, ELASTOGRAPHY AND CAP ASSESSMENT FOR LIVER DISEASE: Training events and feedback representing clinical research programmes in South Africa, Uganda and Kenya, (2024). 10.6084/M9.FIGSHARE.27941958.

[17] B.N. Mayanja, K. Baisley, N. Nalweyiso, F.M. Kibengo, J.O. Mugisha, L. Van der Paal, D. Maher, P. Kaleebu, Using verbal autopsy to assess the prevalence of HIV infection among deaths in the ART period in rural Uganda: a prospective cohort study, 2006-2008, Popul. Health Metr. 9 (2011) 36.

[18] G. O’Hara, J. Mokaya, J.P. Hau, L.O. Downs, A.L. McNaughton, A. Karabarinde, G. Asiki, J. Seeley, P.C. Matthews, R. Newton, Liver function tests and fibrosis scores in a rural population in Africa: a cross-sectional study to estimate the burden of disease and associated risk factors, BMJ Open 10 (2020) e032890.

[19] S. Nsanzimana, M.J. Penkunas, C.Y. Liu, D. Sebuhoro, A. Ngwije, E. Remera, J. Umutesi, C. Ntirenganya, S.D. Mugeni, J. Serumondo, Effectiveness of Direct-acting Antivirals for the Treatment of Chronic Hepatitis C in Rwanda: A Retrospective Study, Clin. Infect. Dis. 73 (2021) e3300–e3307.

[20] J. Umutesi, M.-L. Yu, O. Lesi, J.W. Ward, J. Serumondo, Strategies for Removal of Barriers to Hepatitis C Elimination in Sub-Saharan Africa, J. Infect. Dis. 228 (2023) S221–S225.

[21] B.W. Taye, A Path to Ending Hepatitis C in Ethiopia: A Phased Public Health Approach to Achieve Micro-Elimination, Am. J. Trop. Med. Hyg. 101 (2019) 963–972.

[22] K. Lacombe, R. Moh, C. Chazallon, M. Lemoine, B. Sylla, F. Fadiga, J. Le Carrou, F. Marcellin, C. Kouanfack, L. Ciaffi, M.T. Sartre, M.B. Sida, A. Diallo, J. Gozlan, M. Seydi, V. Cissé, C. Danel, P.M. Girard, T.D. Toni, A. Minga, S. Boyer, P. Carrieri, A. Attia, TAC ANRS12311 Study Group, Feasibility, safety, efficacy and potential scaling-up of sofosbuvir-based HCV treatment in Central and West Africa: (TAC ANRS 12311 trial), Sci. Rep. 14 (2024) 10244.

[23] S.S. Solomon, S. Wagner-Cardoso, L. Smeaton, L.A. Sowah, C. Wimbish, G. Robbins, I. Brates, C. Scello, A. Son, A. Avihingsanon, B. Linas, D. Anthony, E.P. Nunes, D.A. Kliemann, K. Supparatpinyo, C. Kityo, P. Tebas, J.A. Bennet, J. Santana-Bagur, C.A. Benson, M. Van Schalkwyk, N. Cheinquer, S. Naggie, D. Wyles, M. Sulkowski, A minimal monitoring approach for the treatment of hepatitis C virus infection (ACTG A5360 [MINMON]): a phase 4, open-label, single-arm trial, Lancet Gastroenterol Hepatol 7 (2022) 307–317.

[24] Global health estimates, (n.d.). https://www.who.int/data/global-health-estimates (accessed July 6, 2024).

[25] T. Musafiri, I. Kamali, C. Kayihura, J. de la Paix Gakuru, F. Nyirahabihirwe, E. Nizeyimana, P. Kandamage, P. Habinshuti, R. Sekagarura, J.D. Makuza, N. Karema, J. Serumondo, T. Ntakirutimana, J.D. Ndahimana, D.A. Barnhart, Prevalence of hepatitis B and C infection and linkage to care among patients with Non-Communicable Diseases in three rural Rwandan districts: a retrospective cross-sectional study, BMC Infect. Dis. 24 (2024) 247.

[26] J.D. Makuza, C.Y. Liu, C.K. Ntihabose, D. Dushimiyimana, S. Umuraza, M.P. Nisingizwe, J. Umutesi, J. Serumondo, S.D. Mugeni, M. Semakula, N. Gupta, M. Hellard, S. Nsanzimana, Risk factors for viral hepatitis C infection in Rwanda: results from a nationwide screening program, BMC Infect. Dis. 19 (2019) 688.

[27] J.D. Makuza, M.P. Nisingizwe, J.O.T. Rwema, D. Dushimiyimana, D.S. Habimana, S. Umuraza, J. Serumondo, A. Ngwije, M. Semakula, N. Gupta, S. Nsanzimana, N.Z. Janjua, Role of unsafe medical practices and sexual behaviours in the hepatitis B and C syndemic and HIV co-infection in Rwanda: a cross-sectional study, BMJ Open 10 (2020) e036711.

[28] M. de Haas, MOVING BEYOND COLONIAL CONTROL? ECONOMIC FORCES AND SHIFTING MIGRATION FROM RUANDA-URUNDI TO BUGANDA, 1920–60, J. Afr. Hist. 60 (2019) 379–406.

[29] Y.A. Nartey, R. Okine, A. Seake-Kwawu, G. Ghartey, Y.K. Asamoah, A.D.J. Siaw, K. Senya, A. Duah, A. Owusu-Ofori, O. Adarkwa, S. Agyeman, S.A. Bampoh, L. Hiebert, H. Njuguna, N. Gupta, J.W. Ward, L.R. Roberts, A.S. Bockarie, Y.A. Awuku, D. Obiri-Yeboah, Hepatitis C virus seroprevalence, testing, and treatment capacity in public health facilities in Ghana, 2016-2021; A multi-centre cross-sectional study, PLoS One 18 (2023) e0287580.

[30] Liverpool HEP interactions, (n.d.). http://www.hep-druginteractions.org (accessed September 11, 2023).

[31] L. Castéra, J. Vergniol, J. Foucher, B. Le Bail, E. Chanteloup, M. Haaser, M. Darriet, P. Couzigou, V. D. Lédinghen, Prospective comparison of transient elastography, Fibrotest, APRI, and liver biopsy for the assessment of fibrosis in chronic hepatitis C, Gastroenterology 128 (2005) 343–350.

