## Supplementary material for "Micro-elimination of Hepatitis C virus (HCV) infection in the General Population Cohort in rural Uganda: long-term follow-up to assess feasibility and outcomes of a screening and treatment intervention"

**in rural Uganda: feasibility of screening,**

**treatment, and long-term follow-up**

**Contents**

S1: Supplementary methods: assessment before and during HCV treatment	1

S2: Summary table of 13 individuals diagnosed with active HCV infection within the Uganda General Population cohort.	3

S3: Verbal autopsy data recorded locally for six individuals before or after treatment for HCV infection.	4

S4. Sequence data showing amino acid polymorphisms at positions in HCV proteins which are potentially associated with resistance or reduced susceptibility to DAA treatment.	5

References	6

### S1: Supplementary methods: assessment before and during HCV treatment

Any other medications were reviewed to identify potential drug-drug interactions [[1]](https://paperpile.com/c/QfTWA7/cTdEW), although LED/SOF has minimal interactions and is also suitable for treating people who are receiving antiretroviral therapy (ART) for HIV. Exclusion criteria for treatment were (i) significant drug interactions that could not be avoided; (ii) clinically significant illness or any other major medical disorder that could interfere with assessment, treatment, or adherence to the DAA regimen. Individuals were counselled not to initiate any new drug therapies (including traditional remedies) whilst receiving HCV treatment. Prior to treatment, a clinical review was conducted to identify clinical evidence of decompensated liver disease (bleeding from oesophageal or rectal varices, ascites, encephalopathy, jaundice), so that additional assessment and intervention could be implemented.

To support roll-out of a new treatment regimen in this setting, enhanced monitoring was put in place during treatment, with weekly review to assess for side effects and adherence.

Suggested liver stiffness thresholds based on transient elastography in people living with HCV are >7.1 kPa to define F2 fibrosis, >9.5 kPa for F3, and ≥11.0 kPa for F4 (cirrhosis) [[2]](https://paperpile.com/c/QfTWA7/zW7C3).

### S2: Summary table of 13 individuals diagnosed with active HCV infection within the Uganda General Population cohort.

Age, sex and ethnicity redacted to prevent risk of identification. Study flow and three time points are shown in Figure 1.

| **ID** | **HCV-**  **Gt** | **HIV status** | **HBV status** | **Treated for HCV** | **Baseline (2011)** | | **Pre-treatment**  **(2017)** | | | **Follow up**  **(2023)** | | **Status at final follow up** |
| --- | --- | --- | --- | --- | --- | --- | --- | --- | --- | --- | --- | --- |
|  |  |  |  |  | **ALT** | **TE** | **ALT** | **TE** | **HCV RNA VL** | **ALT** | **TE** |  |
| U49 | 4q | Neg | Neg | yes | 24 | 9 | 16 | 6.5 | 1,370,352 | n/a | n/a | Died |
| U100 | 4k | Pos | Neg | yes | 32 | 10.3 | 32 | 13.3 | 6,046,124 | n/a | n/a | Died |
| U149 | 4q | Neg | Neg | no | 71 | n/a | n/a | n/a | n/a | n/a | n/a | Died |
| U150 | 4q | Neg | Neg | yes | 26 | 10.3 | 25 | 9.7 | 23,430 | 8 | 6.8 | Attended review |
| U275 | 4k | Neg | Neg | yes | 26 | 4.4 | 26 | 4 | 235,605 | n/a | n/a | Lost to followup |
| U278 | 4v | Neg | Neg | yes | 25 | 4.4 | 22 | 6.1 | n/a | 5 | 7.1 | Attended review |
| U282 | 4q | Neg | Neg | no | 18 | n/a | n/a | n/a | n/a | n/a | n/a | Died |
| U288 | 7* | Neg | Neg | no | 21 | n/a | n/a | n/a | n/a | n/a | n/a | Lost to followup |
| U294 | 4v | Neg | Neg | no | 10 | n/a | n/a | n/a | n/a | n/a | n/a | Died |
| U295 | 4s | Neg | Neg | yes | 24 | 4.8 | 17 | 6.1 | 7,666,002 | 15 | 4.3 | Attended review |
| U316 | 4v | Neg | Neg | no | 42 | n/a | n/a | n/a | n/a | n/a | n/a | Died |
| U317 | 4k | Neg | Neg | yes | 28 | 8.6 | 11 | 14.1 | 1,471,936 | 18 | 6.5 | Attended review |
| U320 | 4k | Neg | Neg | no | 20 | n/a | n/a | n/a | n/a | n/a | n/a | Died |

ALT - alanine transferase (U/L); Gt - genotype; ID - identifier; TE - transient elastography, measured by Fibroscan (kPa); VL - viral load (IU/L); n/a - not available

* subgenotype not designated

### S3: Verbal autopsy data recorded locally for six individuals before or after treatment for HCV infection.

Methods have been previously reported [[3]](https://paperpile.com/c/QfTWA7/9kDkN)**.** Individual details and identifiers have been redacted to avoid the risk of unintentional disclosure.

| **Recorded history** | **Primary cause of death reported locally** | **Secondary cause of death reported locally** |
| --- | --- | --- |
| **Deaths prior to receipt of HCV treatment** | | |
| Abdominal pain and swelling. Yellow eyes. Swollen feet. Hiccups and became unconscious. | Abdominal problem* | N/A |
| Fever, mental disturbance, swollen body. Abdominal problems. History of bleeding and yellow eyes . Consulted traditional healer and death reported as witchcraft. | Witchcraft* | N/A |
| Known diabetes. Developed abdominal distension and pain, swollen feet. Hospital diagnosis of liver and kidney disease. | Liver disease* | Diabetes |
| Known hypertension. Pain in back and limbs. Weight loss and recent hospitalisation. | Unknown | N/A |
| **Deaths after receiving HCV treatment** | | |
| Painful distended abdomen, hospital diagnosis of liver disease. Nose bleeds. Weight loss. Swollen feet. Got treatment but eventually died. | Liver disease* | N/A |
| Known HIV on treatment. Collapsed and died before reaching hospital.** | Sudden death* | N/A |

N/A not applicable

* Cases marked with an asterisk are those in which liver disease is deemed a likely significant contributor to death based on retrospective clinical review by a senior physician with experience in liver health and infectious diseases.

** This individual had a TE score compatible with cirrhosis.

### **S4. Sequence data showing amino acid polymorphisms at positions in HCV proteins which are potentially associated with resistance or reduced susceptibility to DAA treatment**.

Sequencing methods and interpretation have been previously published [[4]](https://paperpile.com/c/QfTWA7/BS2oQ). First eight rows show genotype-specific reference sequences (‘Refs’) at key positions potentially associated with DAA resistance (wild-type in bold at the top of each column, grouped by the region of the HCV genome). The next 13 rows present HCV sequence data for the individuals represented in this study, where a dash indicates no change from wild-type. Even in the presence of polymorphisms that may be relevant to reduced DAA susceptibility, 100% SVR rate was documented in those who received treatment.

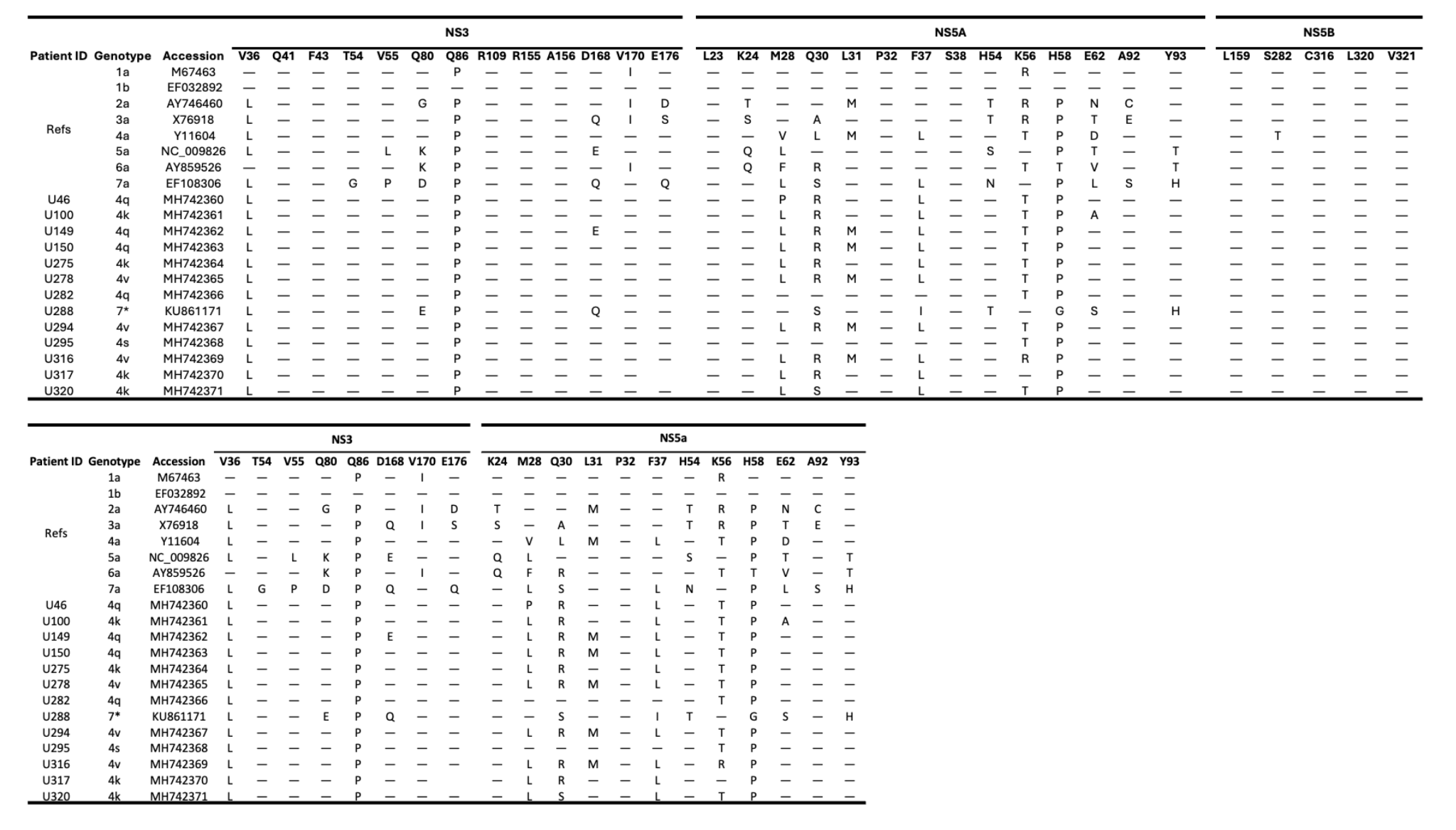

DAA - directing acting antiviral. SVR - sustained virological response (HCV RNA negative at 12+ weeks after treatment completion).

### References

1.	Liverpool HEP interactions. <http://www.hep-druginteractions.org> (accessed 11 September 2023).

2.	**Castéra L, Vergniol J, Foucher J, Le Bail B, Chanteloup E, *et al.*** Prospective comparison of transient elastography, Fibrotest, APRI, and liver biopsy for the assessment of fibrosis in chronic hepatitis C. *Gastroenterology* 2005;128:343–350.

3.	**Mayanja BN, Baisley K, Nalweyiso N, Kibengo FM, Mugisha JO, *et al.*** Using verbal autopsy to assess the prevalence of HIV infection among deaths in the ART period in rural Uganda: a prospective cohort study, 2006-2008. *Popul Health Metr* 2011;9:36.

4.	**Davis C, Mgomella GS, da Silva Filipe A, Frost EH, Giroux G, *et al.*** Highly diverse hepatitis C strains detected in sub-Saharan Africa have unknown susceptibility to direct-acting antiviral treatments. *Hepatology* 2019;69:1426–1441.
